# Threat awareness and counter-terrorism preparedness of Dutch hospitals: a cross-sectional survey

**DOI:** 10.1101/2023.10.14.23297038

**Authors:** Dennis G. Barten, Maud Janssen, Harald De Cauwer, Dennis Keereweer, Edward Tan, Frits van Osch, Luc Mortelmans

**Affiliations:** Department of Emergency Medicine, VieCuri Medical Center, Venlo, the Netherlands; Department of Neurology, Dimpna Regional Hospital, Geel, Belgium; Faculty of Medicine and Health Sciences, University of Antwerp, Wilrijk, Belgium; Department of Trauma Surgery and Emergency Medicine, Radboud University Medical Center, Nijmegen, The Netherlands; Department of Clinical Epidemiology, VieCuri Medical Center, Venlo, the Netherlands; Department of Epidemiology, NUTRIM School for Nutrition Translational Research in Metabolism, Maastricht University, Maastricht, The Netherlands; Center for Research and Education in Emergency Care, University of Leuven, Leuven, Belgium; REGEDIM, Free University Brussels, Brussels, Belgium; Department of Emergency Medicine, ZNA Camp Cadix, Antwerp, Belgium

**Author notes:** **Corresponding author:** D.G. Barten, MD, Department of Emergency Medicine, VieCuri Medical Center, P.O. Box 1926, 5900 BX Venlo, The Netherlands.

## Abstract

**Background:** Workplace violence, including violent extremism, is a growing concern in the healthcare environment. Furthermore, there has been a disproportionate rise in the rate of terrorist attacks on hospitals during the past two decades. Hospitals are vulnerable targets due to their easy accessibility and their high density of patients, staff and visitors. Nonetheless, little is known about the hospitals’ awareness of these risks, and to which extent these facilities protect themselves from violent extremism and terrorist attacks.

**Methods:** This was a cross-sectional survey study among emergency managers of acute care hospitals in the Netherlands. The questionnaire included 42 items across six themes: demographic (hospital) data; general and emergency department (ED) access control; ED security and preparedness; online security and offline transparency; violence, terrorism and warfare; and impact of the COVID-19 pandemic. Responses were collected and stored in a secured online database, and subsequently exported to an Excel spreadsheet for descriptive analysis. Continuous data were reported as means or as medians with interquartile ranges (IQR), using SPSS. Categorical data were reported as absolute numbers and as valid percentages.

**Results:** The questionnaire was completed on behalf of 33 out of 71 hospital organizations (46%), representing 38 out of 82 EDs (46%). Hospitals had broadly different policies with regards to patient and visitor registration, and the presence of security guards. Most hospitals had controlled vehicle access for the parking lot and ambulance bay, but this was not 24/7 in all hospitals. A paragraph on terrorist attacks was included in 34% of hospital disaster plans. Eighteen percent of hospitals had previous experience with acts of violent extremism and 55% of hospitals had sustained (attempted) cyberattacks. Whilst the likelihood of a physical terrorist attack was deemed low at 3.6 (median 4, IQR 2.6) on a 10-point Likert scale, the likelihood of a cyberattack was considered high at 7.3 (median 8, IQR 2.3). A significant proportion of emergency managers reported to experience an increased risk of violence since the onset of the COVID-19 pandemic.

**Conclusion:** Practice variation with regards to counter-terrorism defence measures in Dutch hospitals is high. The preparedness of hospitals for terrorist attacks or acts of violent extremism could be improved and may benefit from uniform, evidence based hospital security policies.

## INTRODUCTION

Workplace violence and violence targeting healthcare constitute a growing concern in the healthcare environment, and the scale of the problem may have been further exacerbated by the anti-science narrative of conspiracy theorists during the COVID-19 pandemic.^1-3^ In addition, it has been shown that healthcare facilities, hospitals in particular, are vulnerable soft targets for terrorist attacks.^4^ In comparison with other targets, there has been a disproportionate rise in the rate of terrorist attacks on hospitals during the past two decades.^5^ Finally, there is a refractory pattern of attacks on healthcare in conflict zones, with 2022 marking the most violent year against health workers and facilities in the last decade.^6,7^

Hospitals are vulnerable targets for acts of violent extremism and terrorism as they must remain easy accessible to the general public and because they are densely occupied with patients and staff 24/7.^4,8^ Furthermore, the intrahospital presence of flammable materials, noxious gasses and radioactive chemicals could cause additional detrimental effects of an attack. A terrorist attack does not only cause acute effects, like high influx of victims, structural damage to the building and hospital staff being injured, it also has long-term effects, including hospital units being unavailable for a long time, the need to replace hospital staff, and staff suffering from post-traumatic stress disorder.^4,5^

Although comparatively rare, secondary attacks may also represent a risk for hospitals. Secondary attacks are deliberate follow-up attacks on hospitals after a primary incident elsewhere. A review of 454 terrorist attacks against hospitals between 1970 and 2019 contained three secondary attacks against hospital emergency departments (EDs). All of these attacks occurred in the past two decades.^4^

Previous studies and reports have suggested several measures to improve hospital security. These include securing and guarding points of entries, limiting access with ID badges or biometrics, utilization of metal detectors, vehicle and personnel entry screening, armed security guards, and closed-circuit television (CCTV).^9^ However, little is known about the efficacy of these precautions, and to which extent they have been adopted by hospitals.

The aim of this cross-sectional survey was to assess the threat awareness of hospitals in the Netherlands, and to which extent these facilities adopted security measures to protect themselves from violent extremism and terrorist attacks.

## METHODS

### Study design

This was a cross-sectional survey using questionnaires among emergency managers of all acute care hospitals (hospitals including an ED and Intensive Care unit (ICU)) in the Netherlands. Emergency managers were invited by email (directly, via general contact addresses of the hospitals, or through the liaison officers of one of the Dutch acute care networks) and were sent reminder emails after two and six weeks. The survey was conducted between April and June 2022.

The questionnaire (Appendix I) was designed by the research team and was based on literature research and expert consultation. It included 42 items across six themes: demographic (hospital) data; general and ED access control; ED security and preparedness; online security and offline transparency; violence, terrorism and warfare; and impact of the COVID-19 pandemic. Informed consent was obtained from all participants in the first item of the questionnaire.

The majority of questions was multiple choice. For some questions, respondents were able to provide additional information in a blank space when choosing the option ‘other:’.

Furthermore, some questions included a ranking on a 10-point Likert scale. The questionnaire was created using Castor EDC (Amsterdam, The Netherlands), a secured online data collection programme. Hospital organisations with >1 ED, had to complete the survey for every ED. The results were processed anonymously but were not blinded (to the researchers) with regards to the respective hospitals.

### Setting

The Netherlands has a modern healthcare system with effective primary care and specialised acute and critical care facilities. In 2021, there were 82 hospital-based EDs under the umbrella of 71 different hospital organizations.^10^ Dutch law requires all hospitals to have a hospital disaster preparedness plan (HDPP), but requirements regarding the contents or design are not specified.^11^ Resultantly, a chapter on violence, extremism and terrorism is not mandatory. The Netherlands ranks into the medium zone of the Global Terrorism Index, with a total of 137 terrorist attacks between 1970 and 2021.^12^

### Statistics

All data were exported into an Excel spreadsheet (Microsoft Corp.; Redmond, Washington, USA) for analysis. Descriptive analyses were performed. Continuous data were reported as means, or as medians with interquartile ranges (IQR) if continuous variables were not normally distributed, using SPSS (SPSS Inc., Chicago, USA). Categorical data were reported as absolute numbers and as valid percentages.

### Ethical approval

Ethical approval of this study was waived by the medical-ethical review board of Maastricht University Medical Center based on Dutch legislation for research concerning humans (2021-2548; Maastricht, the Netherlands).

## RESULTS

The questionnaire was completed by 33 out of 71 hospital organisations (46%), representing 38 out of 82 EDs (46%). In seven of the questionnaires there were one or more missing fields.

The participating hospitals were equally distributed over country. The number of beds per hospital varied from 140 to 980. The annual ED census varied from 7,500 to 50,000 (mean: 35,000). The baseline characteristics of the hospital organizations are shown in Table 1.

**Table 1.** Baseline characteristics hospital organizations*.

| **Characteristic** | **Hospital organizations (n=33)** |
| --- | --- |
| Academic | 3 (9%) |
| Teaching hospital | 16 (48%) |
| Peripheral | 14 (42%) |
| 1 ED | 28 (85%) |
| 2 EDs | 5 (15%) |
| Urban | 31 (94%) |
| Rural | 2 (6%) |

\*Hospital organizations may represent multiple hospital (ED) locations

### General and ED access control

Thirty-three hospitals (87%) had a registration desk for patients at the main hospital entrance. None of the hospitals registered the entrance or presence of visitors. In 35 hospitals (92%), there was entrance control of staff members using ID badges for specific staff entrances and certain areas in the hospital.

Twenty-seven hospitals (71%) reported to have in-hospital security personnel at the main entrance and/or ED. Two hospitals (5%) used a metal detector at the main entrance. Weapon checks on patients and visitors in the ED were always performed in four hospitals (11%), and only during crisis situations in another nine hospitals (24%). These situations were not further specified.

Video surveillance (CCTV) was available in 36 hospitals (95%). Thirty hospitals (79%) used video surveillance 24/7; two hospitals reported that they did not actively monitor the CCTV.

Vehicle access to the ED parking lot and ambulance bay was gate-controlled 24/7 in 27 hospitals (71%), in 1 hospital (3%) during business hours only, and in nine hospitals (24%) during out-of-office hours only. In 26 hospitals (68%), only licensed ambulances were granted access to the ambulance bay. In seven hospitals (18%), both ambulances and non-urgent patient transport vehicles had access to the ambulance bay, and ambulance bay access was unrestricted in five hospitals (13%).

### ED security and preparedness

Twenty-three hospitals (61%) had a template protocol and/or agreements with law enforcement in order to safeguard the ED in case of incidents. Eight EDs (21%) had an alternative ED location at their disposal, where the operational activities of the ED can be managed in case of an incident at the original ED. Another seven hospitals (18%) had a control centre (at another location than the ED) for incident management.

Eleven hospitals (29%) were able to manage patients with nuclear contamination 24/7. Another nine hospitals (24%) could only deal with nuclear contamination when an expert of the nuclear medicine department was present, and nine hospitals had delegated this service to external parties, mostly the fire brigade. Four hospitals (11%) were unable to respond to nuclear emergencies.

A decontamination unit was available in 32 hospital EDs (84%). Of these, 23 (61%) were located outside the ED, 7 (18%) at the entrance of the ED and 2 (5%) within the ED. In 30 out of the 32 hospitals with a decontamination unit, ED staff members received specific training to adequately respond to chemical, biological, radiological and nuclear (CBRN) threats.

### Online security and offline transparency

Twenty-five hospitals (66%) listed the names (and photos) of board members and/or staff members on their homepage. Twenty-four hospitals (63%) did have one or more of the following online accessible: maps, virtual tours, photos of the hospital, and vlogs. One hospital (3%) did not have any of the abovementioned online, and another hospital only had this information available in their secured online patient platform.

Seven hospitals (18%) reported to organize tours for the general public on a regular basis, 11 hospitals (29%) organized such tours for students only, and six hospitals (16%) reported to have specific tours for police and fire department, for new employees, or to prepare patients before surgery. Eight hospitals (21%) organized yearly open days instead of hospital tours. There were no such activities in six hospitals.

### Violence, terrorism and warfare

All hospitals had a HDPP and all hospital organized disaster drills, which were performed periodically in 30 hospitals (79%) and for new staff members only in eight hospitals (21%). In 13 hospitals (34%), the HDPP included a paragraph about extreme violence or terrorism directly targeting the hospital. Of these, five hospitals (13%) also had an emergency plan in case of a secondary attack. In case of a terrorist attack in the vicinity of the hospital, seven hospitals (18%) would go in lockdown, around six hospitals (16%) checkpoints and road blocks would be installed, and one hospital (3%) would be provided with police surveillance. No single hospital reported to have a bomb shelter where patients and staff can hide in case of an emergency.

Seven hospitals (18%) previously experienced acts of violent extremism or terrorism. These included threats from criminals (n=2), incidents with firearms (n=1) or stabbing weapons (n=1), a bomb threat in the city centre, riots, bullets destroying a car in the ambulance bay, and a COVID-19 protest which culminated in extreme violence targeting the hospital. All seven hospitals were urban, but no further associations were found between hospital type and the exposure to such violent incidents. Twenty-one hospitals (55%) reported an attempted cyberattack. In two hospitals, a cyberattack resulted in patient data leaks and the interruption of patient care.

On a Likert scale of 0-10 (0 = no threat, 10 = serious threat), emergency managers rated the likelihood of a physical terrorist attack in their hospital 3.6 (median 4, IQR 2.58). The likelihood of a cyberattack was rated 7.3 (median 8, IQR 2.31).

### Impact of the COVID-19 pandemic

Twelve hospitals (32%) experienced more threats since the COVID-19 pandemic, aggressive behaviour by patients and visitors in particular. During the pandemic, access control was enhanced in 29 hospitals (76%): selected entrances were closed, (additional) security guards were deployed, and security protocols were re-evaluated.

Eight hospitals (21%) reported that there was less attention to the threat of terrorist incidents and violent extremism because of the pandemic. Emergency managers explained that they were focused on managing the pandemic, and lacked time for counter-terrorism training. Fifteen hospitals (39%) denied to have spent less attention to these threats. In fact, they reported that there were more security measures in place, and that threat awareness of hospitals was already low before the pandemic.

## DISCUSSION

This nationwide questionnaire-based study aimed to provide insight into the threat awareness and preparedness of Dutch hospitals for terrorist attacks or acts of violent extremism. It was shown that there is considerable practice variation between hospitals with regards to access control and security measures. Although emergency managers deemed the likelihood of a physical terrorist attack in their hospital low, one third of the hospitals experienced more threats since the COVID-19 pandemic, while this same pandemic also distracted from the focus on other crises. Furthermore, the majority of hospitals have already experienced cyberattacks.

Within hospitals, the ED environment is at highest risk of violent incidents. Of all shootings in US hospitals, the ED was the most common site (29%), followed by the parking lot (23%) and patient rooms (19%).^13,14^ A 2008 survey on workplace violence in US EDs revealed that guns or knives were brought to the ED on a daily or weekly basis in 20% of the hospitals.^15^ Nonetheless, no standard exists regarding weapons screening in the ED. As a result, the majority of US EDs operate with a complete lack of weapons screening.^14^ In the Netherlands, gun violence is uncommon and hospital shootings have incidentally occurred. However, the recent combined arson attack and hospital shooting in Erasmus University Medical Center (Rotterdam, September 2023), killing one healthcare professional, has demonstrated that the risk is not negligible.^16^ Hospitals should consider to review their access policy, and they may differentiate between general hospital access and ED access, as the latter is more prone to violent attacks. However, strict access control and improved security will not prevent all violent incidents. It may therefore be helpful to train clinical and non-clinical staff for such threatening situations, including active shooter scenarios.^17,18^ In all Dutch hospitals, access control of hospital was focused on patients and staff, but not on visitors. Furthermore, there was significant practice variation regarding the presence of security personnel and the execution of security checks. Video surveillance was present in most hospitals, but this was not actively monitored 24/7 in all facilities. This suggests that there are several opportunities for potential perpetrators to enter the hospital without being noticed.

Vehicle access to the ED parking lot and ambulance bay was controlled in the majority of hospitals. However, ambulance bay access was not limited to ambulances only in all hospitals. In a previous study, the ambulance bay was identified as a potential window of opportunity for terrorists, because ambulances are often left unlocked and unattended in the ambulance bay. As a result, these ambulances are susceptible for possible hijacking purposes.^19^ Two other studies showed that ambulances are increasingly used by terrorists to conduct terrorist attacks. When there is little or no access control of the ambulance bay, this may pose a security risk for both the hospital and the community.^20,21^

The online and offline transparency of hospitals is increasing, which is positive but may also increase their vulnerability. Perpetrators may use available information online, including names and photos of staff members, maps, virtual tours and photos of the hospital, to prepare an attack. Hospital organisations should therefore critically appraise their online presence for possible security risks and vulnerabilities.^3,8^

Although all hospitals had a HDP in place and most hospitals performed periodical disaster drills, they often lacked a chapter about extreme violence or terrorism. Likewise, a recent study that assessed the content and quality of Dutch HDPPs found that the safety of Dutch hospitals during the disaster and post-disaster recovery phase is suboptimal according to World Health Organization (WHO) checklists.^11^ In this present study, 1 in 5 hospitals reported to have experienced acts of violent extremism. Although there is a thin line between violent extremism and terrorism, no documented terrorist attacks against hospitals in the Netherlands have occurred to date - unless the previously mentioned Rotterdam hospital shooting will be classified as terrorism. Still, it should be noted that the risk of terrorist attacks against hospitals does exist. Between 1970-2019, there were 454 attacks against hospitals worldwide, of which 29 took place in Europe.^4^ Another concern is the reported increase of attacks on healthcare in armed conflicts, which is a daily occurrence in in many conflict zones, including Ukraine.^6^ Furthermore, hospitals experienced more threats of violence since the COVID-19 pandemic and many hospitals had to intensify their security measures.^2^ Nonetheless, emergency managers deemed the likelihood of a physical terrorist attack in their hospital relatively low. With more than half of the hospitals having been the victim of cybercrimes, the risk of a cyberattack was deemed significantly higher. Although it is unknown whether this also represents better preparedness, several studies have shown that cybersecurity in hospitals can be significantly improved.^22,23^

### Limitations

There are limitations to this study. First, non-responder bias could exist, although the response rate of 46% is reasonable and the baseline characteristics are likely representative for the Dutch hospital landscape. Second, while all hospitals had emergency managers, their role differed considerably. In some cases, they could not answer all questions and had to consult other hospital staff members, mostly emergency physicians. This could have impacted the results, as emergency physicians may have another perspective and training than emergency managers. At the same time, this may represent the reality of crisis management in hospitals, which is not limited to one liaison. Third, some questions related to past experiences, which may be prone to recall bias. Furthermore, it was noted that two hospitals refused to answer these questions. A strength of this study is that it is one of the first to address this specific topic. Although this study is geographically limited to the Netherlands, the results may be representative for other Western European countries, which have comparable similar healthcare systems and threat environments. However, further region- and country-specific studies are warranted. Future studies should assess best practices and the feasibility and efficacy of specific security measures.

## CONCLUSION

Practice variation with regards to counter-terrorism defence measures in Dutch hospitals is high. The preparedness of hospitals for terrorist attacks or acts of violent extremism could be improved and may benefit from uniform, evidence based hospital security policies.

## Data Availability

All data produced in the present work are contained in the manuscript

## ABBREVIATIONS

CBRN: chemical, biological, radiological and nuclear
CCTV: closed-circuit television
ED: emergency department
GTD: Global terrorism database
HDPP: hospital disaster preparedness plan
ICU: intensive care unit
IQR: interquartile ranges
WHO: World Health Organization

## APPENDIX I questionnaire

### Demographic section

Name of hospital organization:

City/town:

Bed capacity (per location):

Number of emergency departments (EDs):

Annual ED census (per ED):

Hospital type: academic, teaching or peripheral

### A. General and ED access control

1. Is the entrance of patients or visitors registered?
  - Yes, patients only
  - Yes, visitors only
  - Yes, both patients and visitors
  - No
2. Is there access control of staff?
  - Yes, with ID badges
  - Yes, with biometrical identification
  - No
  - Other (please specify):
3. Is vehicle access to the ED parking lot and ambulance bay gate-controlled?
  - Yes, 24 hours/day
  - Yes, during office hours only
  - Yes, out-of-office hours only
  - No
4. Is the hospital situated in an urban or rural location?
  - Urban
  - Rural
5. Who has access to the ambulance bay?
  - Licensed ambulances only
  - Ambulances and non-urgent patient transport vehicles
  - Unrestricted access
6. Are metal detectors deployed at the entrance?
  - Yes, at the main entrance only
  - Yes, at the ED only
  - Yes, both main entrance and ED
  - No
  - Other (please specify):
7. Are bag checks performed?
  - Yes, at the main entrance only
  - Yes, at the ED only
  - Yes, both main entrance and ED
  - No
  - Other (please specify):
8. Are weapon checks performed on patients and/or visitors in the ED?
  - Yes, all patients (including those brought in by ambulance)
  - Yes, except for patients who are brought in by ambulance
  - In crisis situations only
  - No
9. Does in-hospital security personel monitor access control?
  - Yes, at the main entrance only
  - Yes, at the ED only
  - Yes, both main entrance and ED
  - No
  - Other (please specify):
10. When does in-hospital security personel monitor access control? o 24/7
  - During office-hours
  - Out-of-hours
  - Never
  - Other (please specify):
11. Is video surveillance (CCTV) available in your hospital?
  - Yes
  - No
12. When is CCTV monitored?
  - 24/7
  - During office-hours
  - Out-of-hours
  - Never
  - Other (please specify):

### B. ED: security and preparedness

13. Is there an escape room where staff can be put in safety in case of incidents in the ED?
  - Yes
  - No
14. Is there a template protocol with law enforcement in order to safeguard the ED in case of incidents?
  - Yes, there is an emergency button/alarm with direct activation of the police
  - Yes, there is permanent ED guarding by the police
  - No, there is only police guarding with a local terrorist threat
  - No, there is only police guarding with a national terrorist threat
  - No
  - Other (please specify):
15. Does the hospital have an alternative ED location and/or emergency control center at its disposal?
  - Yes, there is an alternative ED location
  - Yes, there is an emergency control center
  - No
16. Are the following items available at the ED?
  - Tourniquets
  - Combat gauzes and/or bleeding control kits
  - Disaster kit
  - Tranexamic acid
  - None of the above
  - Other (please specify):
17. Are the items in Q16 available at the other hospital location?
  - Yes (please specify items in Q16)
  - No
18. Is your hospital able to manage patients with nuclear contamination?
  - Yes, 24/7
  - Yes, only when an expert of the nuclear medicine department is present
  - Yes, through external parties (i.e. fire brigade)
  - No
  - Other (please specify):
19. Is there a decontamination unit available at the ED?
  - Yes
  - No (continue with Q21)
20. Where is the decontamination unit situated?
  - Outside the ED
  - At the ED entrance
  - In the ED
21. Is the staff trained in the use of the decontamination unit?
  - Yes
  - No
22. Is there sufficient personal protective equipment (PPE) for CBRN incidents?
  - Yes
  - No (continue with Q24)
23. Is the staff sufficiently trained in the use of PPE in case of CBRN incidents?
  - Yes
  - No

### C. Online security and offline transparency

24. Does the hospital’s website or social media display one of the following items? (multiple items may apply):
  - Map of the hospital
  - Map of the ED
  - Virtual hospital tour
  - Virtual ED tour
  - Photos of affiliated ambulances
  - Vlogs
  - None of the above
  - Other (please specify):
25. Does the hospital’s website or social media display one of the following items? (multiple items may apply):
  - Names and/or photos or board members
  - Names and/or photos of physicians
  - Names and/or photos of nurses
  - No
  - Other (please specify):
26. Does the hospital organize tours for the general public?
  - Only during open days
  - For students only
  - Regularly, for various types of public
  - No
  - Other (please specify):
27. Does your hospital previously experience a cyberattack?
  - Yes, failed attack
  - Yes, with patient data leak
  - Yes, with failure of electronic health records
  - Yes, with consequences for patient care
  - No
28. Is the hospital affiliated with a cybersecurity service to manage (the consequences of) cyberattacks?
  - Yes, internally 24/7
  - Yes, externally (specialized company)
  - Ja, internally during office-hours
  - No
  - Other (please specify):

### D. Violence, terrorism and warfare

29. Does the HDPP include a protocol in case of a terrorist threat in the vicinity of the hospital? (multiple answers may apply):
  - Yes, the hospital will be guarded by police
  - Yes, the hospital will go in lockdown
  - Yes, road blocks and/or checkpoints will be installed around the hospital
  - No
  - Other (please specify):
30. Is all staff familiar with the content of the hospital disaster preparedness plan?
  - Yes
  - No
31. Are disaster scenario drills performed in your hospital?
  - Yes, for new staff members only
  - Yes, periodically
  - No
32. Does your hospital have experience with extreme violence or terrorism?
  - Yes (please specify):
  - No
33. Does the HDPP include a chapter on extreme violence or terrorism targeting the hospital?
  - Yes
  - No (continue with Q34)
34. Is all staff familiar with this specific chapter?
  - Yes
  - No
35. Does the HDPP include a chapter on a secondary terrorist attack* on the hospital?
  - Ja, namelijk
  - Nee *Secondary attacks are deliberate follow-up attacks on hospitals after a primary incident elsewhere.
36. Is there a bombshelter in your hospital that can be used in case of military or nuclear threats?
  - Yes
  - No

### E. Impact of the COVID-19 pandemic

37. Did the threat of terrorism or extreme violence for hospitals increase since the COVID-19 pandemic?
  - Yes
  - No
38. On a 0-10 Likert scale, how would you score the likelihood of a physical terrorist attack on your hospital? (0 = no threat, 10 = severe threat)
  - 0 1 2 3 4 5 6 7 8 9 10
39. On a 0-10 Likert scale, how would you score the likelihood of a cyberattack on your hospital? (0 = no threat, 10 = severe threat)
  - 0 1 2 3 4 5 6 7 8 9 10
40. Did the COVID-19 pandemic change access control?
  - Yes, enhanced access control
  - Yes, diminished access control
  - No
41. Did the pandemic lower the attention towards prevention of terrorism or extreme violence?
  - Yes (please specify):
  - No
42. During the pandemic, were there additional security measures in place in your hospital?
  - Yes (please specify):
  - No

## Notes

**Authors’ disclosures** The authors attest the original nature of the material and not to have any financial or other relationships that could be construed as a conflict of interest.

### Competing Interest Statement

The authors have declared no competing interest.

### Funding Statement

This study did not receive any funding

## REFERENCES

1. Rossi MF, Beccia F, Cittadini F, et al. Workplace violence against healthcare workers: an umbrella review of systematic reviews and meta-analyses. Public Health 2023; 221: 50–9.

2. van Stekelenburg BCA, De Cauwer H, Barten DG, Mortelmans LJ. Attacks on Health Care Workers in Historical Pandemics and COVID-19. Disaster Med Public Health Prep 2022; 17: e309.

3. De Cauwer HG, Somville F. Health Care Organizations: Soft Target during COVID-19 Pandemic. Prehosp Disaster Med 2021; 36(3): 344–7.

4. Ulmer N, Barten DG, De Cauwer H, et al. Terrorist Attacks against Hospitals: World-Wide Trends and Attack Types. Prehosp Disaster Med 2022; 37(1): 25–32.

5. McNeilly B, Jasani G, Cavaliere G, Alfalasi R, Lawner B. The Rising Threat of Terrorist Attacks Against Hospitals. Prehosp Disaster Med 2022; 37(2): 223–9.

6. Barten DG, Tin D, Granholm F, Rusnak D, Van Osch F, Ciottone G. Attacks on Ukrainian healthcare facilities during the first year of the full-scale Russian invasion of Ukraine. Medrxiv 2023.

7. Coalition SHiC. Ignoring red lines: violence against health care in conflict, 2022.

8. De Cauwer H, Somville F, Sabbe M, Mortelmans LJ. Hospitals: Soft Target for Terrorism? Prehosp Disaster Med 2017; 32(1): 94–100.

9. Tin D, Hart A, Ciottone GR. Hardening hospital defences as a counter-terrorism medicine measure. Am J Emerg Med 2021; 45: 667–8.

10. O’Connor RD, Barten DG, Latten GHP. Preparations of Dutch emergency departments for the COVID-19 pandemic: A questionnaire-based study. PLoS One 2021; 16(9): e0256982.

11. Blanchette RADGE, Van Bree EM, Bierens JLM. Hospital disaster preparedness in the Netherlands. Int J Disaster Risk Red 2023; 93.

12. Tin D, Barten DG, De Cauwer H, Mortelmans LJ, Ciottone GR. Terrorist Attacks in Western Europe: A Counter-Terrorism Medicine Analysis. Prehosp Disaster Med 2022; 37(1): 19–24.

13. Wax JR, Cartin A, Craig WY, Pinette MG. U.S. acute care hospital shootings, 2012-2016: A content analysis study. Work 2019; 64(1): 77–83.

14. Jasani G, Gaddis GM. Weapons Screening in the Emergency Department: What Is the Standard? J Emerg Med 2021; 60(5): 677–8.

15. Kansagra SM, Rao SR, Sullivan AF, et al. A survey of workplace violence across 65 U.S. emergency departments. Acad Emerg Med 2008; 15(12): 1268–74.

16. Sauer P. Rotterdam shootings: three killed including girl, 14, with man arrested. The Guardian. 2023 29 September 2023.

17. Chu JL, Castaldi M, Bridges K, et al. Besieged in the Bronx: Lessons from an In-Hospital Mass Casualty. J Trauma Acute Care Surg 2023.

18. Regan EM, Cranmer T, Hanaway T. Multimodal Active Shooter Training for Emergency Department Personnel: An Initiative for Knowledge, Comfort, and Retention. Disaster Med Public Health Prep 2021; 17: e63.

19. Alves DW, Bissell RA. Ambulance snatching: how vulnerable are we? J Emerg Med 2003; 25(2): 211–4.

20. Jasani GN, Alfalasi R, Cavaliere GA, Ciottone GR, Lawner BJ. Terrorists Use of Ambulances for Terror Attacks: A Review. Prehosp Disaster Med 2021; 36(1): 14–7.

21. Besenyo J, Barten DG, De Cauwer HG, Tin D, Gulyas A. A Review of Ambulance Terrorism on the African Continent. Prehosp Disaster Med 2023; 38(2): 237–42.

22. van Boven LS, Kusters RWJ, Tin D, et al. Hacking Acute Care: A Qualitative Study on the Health Care Impacts of Ransomware Attacks Against Hospitals. Ann Emerg Med 2023.

23. Kruse CS, Frederick B, Jacobson T, Monticone DK. Cybersecurity in healthcare: A systematic review of modern threats and trends. Technol Health Care 2017; 25(1): 1–10.

